# Clinical Decision Support in Cardiovascular Medicine: Effectiveness, Implementation Barriers, and Regulation

**DOI:** 10.1101/2021.02.26.21252556

**Authors:** Yuan Lu, Edward R. Melnick, Harlan M. Krumholz

## Abstract

Despite considerable progress in addressing cardiovascular disease (CVD) over the past 50 years, there remain many gaps in CVD care quality. Multiple missed opportunities have been identified at every step in the prevention and treatment of CVD, such as failure to make risk factor modifications, failure to diagnose CVD, and failure to use proper evidence-based treatments. With the digital transformation of medicine and advances in health information technology, clinical decision support (CDS) tools offer promise to enhance the efficiency and effectiveness of cardiovascular care delivery. Yet, to date, the promise of CDS delivering scalable and sustained value for patient care in clinical practice has not been realized. Here, we review evidence on key emerging questions around the development, implementation, and regulation of CDS with a focus on CVD. We first review evidence on the effectiveness of CDS on patient health and health delivery outcomes related to CVD and features predictive of effectiveness. We then review the barriers encountered during CDS implementation in cardiovascular care with a focus on unintended consequences and strategies to promote successful implementation. Finally, we review the current legal and regulatory environment of CDS with specific examples for CVD.

## Introduction

Despite tremendous progress in addressing cardiovascular disease (CVD) over the past 50 years, there remain many gaps in CVD care quality. Multiple missed opportunities have been identified at every step in the prevention and treatment of CVD, such as failure to make risk factor modifications, failure to diagnose CVD, and failure to use proper evidence-based treatments.^1^ Further, the value of cardiovascular care in the United States (US) is declining as CVD healthcare costs are continuing to rise and previous decreases in population CVD mortality are slowing.^2^ Consequently, cardiovascular care remains *both* expensive and far short of an evidence-based ideal standard of care.

With the digital transformation of medicine and advances in health information technology, clinical decision support (CDS) tools offer great promise to enhance efficiency and effectiveness of cardiovascular care delivery. The uptake of electronic health record (EHR) systems in hospitals and office practice settings provides opportunities to collect patient-level clinical information efficiently.^3 4^ CDS tools integrated with the EHR can analyze patient data and trigger timely, actionable, evidence-based recommendations to health care teams to support their care decisions.^5^

Yet, to date, the promise of CDS delivering scalable and sustained value for patient care in clinical practice has not been realized. Prior systematic reviews that are not specific to CVD have found only a modest effect of CDS with process of care improvements that are less than 5%.^6–18^ Often CDS interventions intend to provide valuable information to clinicians but result in other unintended consequences when implemented in a real clinical environment.^19^ Additionally, the regulatory environment of CDS is changing - government agencies such as the US Food and Drug Administration (FDA) have released new policies to provide more streamlined oversight for CDS,^20^ which has important implications for promoting widespread adoption and implementation of CDS. Understanding emerging issues surrounding the development, implementation, and regulation of CDS is critical to optimize the meaningful use of CDS in cardiovascular care.

Accordingly, the objectives of this review are (1) to provide a contemporary assessment of the effectiveness of CDS on a wide variety of health care process and clinical outcomes related to CVD; (2) to review the barriers encountered during CDS implementation in cardiovascular care with a focus on the unintended consequences and strategies to promote successful implementation; and (3) to review the current legal and regulatory environment for CDS implementation with specific examples for CVD.

### Incidence and prevalence

CVDs are a group of heart and blood vessel disorders including coronary artery disease, stroke, heart failure, peripheral vascular disease, and other conditions. CVD is the leading cause of mortality and morbidity globally, accounting for 18.6 million deaths per year, a number that is projected to grow to more than 23.6 million by 2030.^21^ According to the Global Burden of Disease Study, there were an estimated 523 million prevalent cases of CVD (6,431 cases per 100,000 persons) in 2019.^21 22^ The age-standardized prevalence of CVD varied significantly by country, ranging from less than 5,000 cases per 100,000 persons in countries in Western Europe, North America, and the Asian Pacific region to more than 9,000 cases per 100,000 persons in countries in West Africa. From 2010-2019, the prevalence of CVD has increased for almost all non-high-income countries. An additional concern is that the prevalence of CVD has also begun to rise in some locations where it was previously declining in high-income countries. The incidence of CVD worldwide was 55.5 million or 684.3 cases per 100,000 persons in 2019. With an aging population and population growth, the number of adults affected by CVD worldwide is expected to continue to rise in future years.

### Sources and selection criteria

We identified sources through a search of PubMed for English language articles from its inception to 8 July 2020, using the concepts and search terms presented in **Table 1**. Concepts were combined using the Boolean and Proximity operator ‘AND’, while search terms within each concept were combined using ‘OR’. Our inclusion criteria were any observational studies or randomized controlled trials (RCTs) that implemented CDS in a real clinical setting for use by health care providers to aid decision making at the point of care for cardiovascular care. We also searched the reference lists of relevant systematic reviews and meta-analyses and included additional selected observational studies and RCTs from these sources. We excluded studies that described nonelectronic CDS, included fewer than 50 participants, described patient-facing decision aids that did not involve a provider, were not relevant to cardiovascular conditions, did not quantify the effect of CDS, did not contain primary study data analyses such as study protocols and opinion pieces, and were not published in English.

**Table 1.** Key words used in search strategies.

| Concept* | Relevant key words |
| --- | --- |
| Clinical decision support | "decision-support"[Title/Abstract] OR "decision-support"[Title/Abstract] OR "clinical support system"[Title/Abstract] OR ("evidence-based"[Title/Abstract] AND "support system"[Title/Abstract]) OR "Decision Support Systems, Clinical"[Mesh] |
| Cardiovascular medicine | cardi*[Title/Abstract] OR coronar*[Title/Abstract] OR heart*[Title/Abstract] OR myocardi*[Title/Abstract] OR hypertension[Title/Abstract] OR endocardi*[Title/Abstract] OR aneurysm*[Title/Abstract] OR hypertension[Title/Abstract] OR hypertensive[Title/Abstract] OR embolism*[Title/Abstract] OR thrombosis[Title/Abstract] OR thromboses[Title/Abstract] OR hypotension[Title/Abstract] OR hypotensive[Title/Abstract] OR stroke*[Title/Abstract] OR "Cardiology"[Mesh] OR "Cardiology Service, Hospital"[Mesh] OR "Cardiovascular Diseases"[Mesh] |
| Study type | clinicalstudy[Filter] OR clinicaltrial[Filter] OR controlledclinicaltrial[Filter] OR evaluationstudies[Filter] OR meta-analysis[Filter] OR multicenterstudy[Filter] OR observationalstudy[Filter] OR pragmaticclinicaltrial[Filter] OR randomizedcontrolledtrial[Filter] OR systematicreview[Filter] |

\*Concepts were combined using the Boolean & Proximity operators “AND”, and the search terms within each concept were combined with “OR”.

For each included study, we extracted the following information into a standardized abstraction spreadsheet: study design, country in which the study conducted, time period, study setting, sample size, clinical domain of the intervention, intervention and control specifications, study outcomes, unintended consequences reported, user experience reported, barriers to CDS implementation reported, and CDS features associated with effectiveness. Given that studies included in this review compared a wide range of interventions for multiple outcomes, we synthesized the data qualitatively to describe the effects of CDS and identify barriers encountered during implementation. For each subsequent section of the paper, we focused the discussion on evidence from large RCTs when available and including the results of smaller RCTs and observational studies when of particular interest or when other evidence was unavailable. For the CDS regulation section, we identified and reviewed regulatory documents from government agencies in the US and European Union (EU).

### Overview of studies of CDS interventions in cardiovascular care

We screened 392 unique references from our search and identified 77 studies that met all inclusion criteria (**Figure 1**). Of the 77 studies that evaluated CDS interventions in cardiovascular care, 2 (2.6%) were published before 2000, 25 (32.5%) in 2000-2010 and 50 (64.9%) after 2010 (**eTable 1 in the Supplement**). There were 34 (44.1%) studies conducted in North America, 28 (36.4%) in Europe, 8 (10.4%) in Asia, 6 (7.8%) in Oceania, and 1 (1.3%) in South America. Most studies (58; 75.6%) used a randomized design, with 40 (51.3%) using a clustered randomized design, allocating intervention status to clinics or provider groups rather than patients. Most studies (59; 75.3%) took place in outpatient/ambulatory settings, while 18 (23.4%) studies occurred in emergency department/inpatient settings. The duration of study was <= 12 months for 33 (42.9%) studies, >12-24 months for 29 (37.7%) studies, and >24 months for 15 (19.5%) studies. The clinical areas most commonly targeted were management (30 studies), preventive care (29 studies), diagnosis (10 studies), and screening (8 studies). The most common types of CDS included: (1) tailored reminders for screening for CVD risk factors, clinical tests, and treatments; (2) risk assessments for developing CVD based on patients’ demographic and clinical risk factors; (3) order sets for initiation and intensification of evidence-based treatments for CVD; (4) patient behavior change recommendations for CVD, such as quitting smoking and increasing physical activity; and (5) alerts when indicators for CVD risk factors are not at goal. While most studies assessed CDS in isolation for cardiovascular care, 13 studies (16.9%) employed CDS within a multicomponent approach to address clinical practice changes targeted at the patient, provider, organizational, or community levels. These approaches ranged from organizational change such as team-based care in which clinicians worked together with pharmacists and nurses to improve healthcare delivery to combining CDS with other implementation strategies such as patient reminders. Twelve (15.6%) studies were conducted in a single site versus 65 (84.4%) studies that were conducted in two or more sites. Only one study validated CDS tools in a separate population.^23^

**Figure 1.**
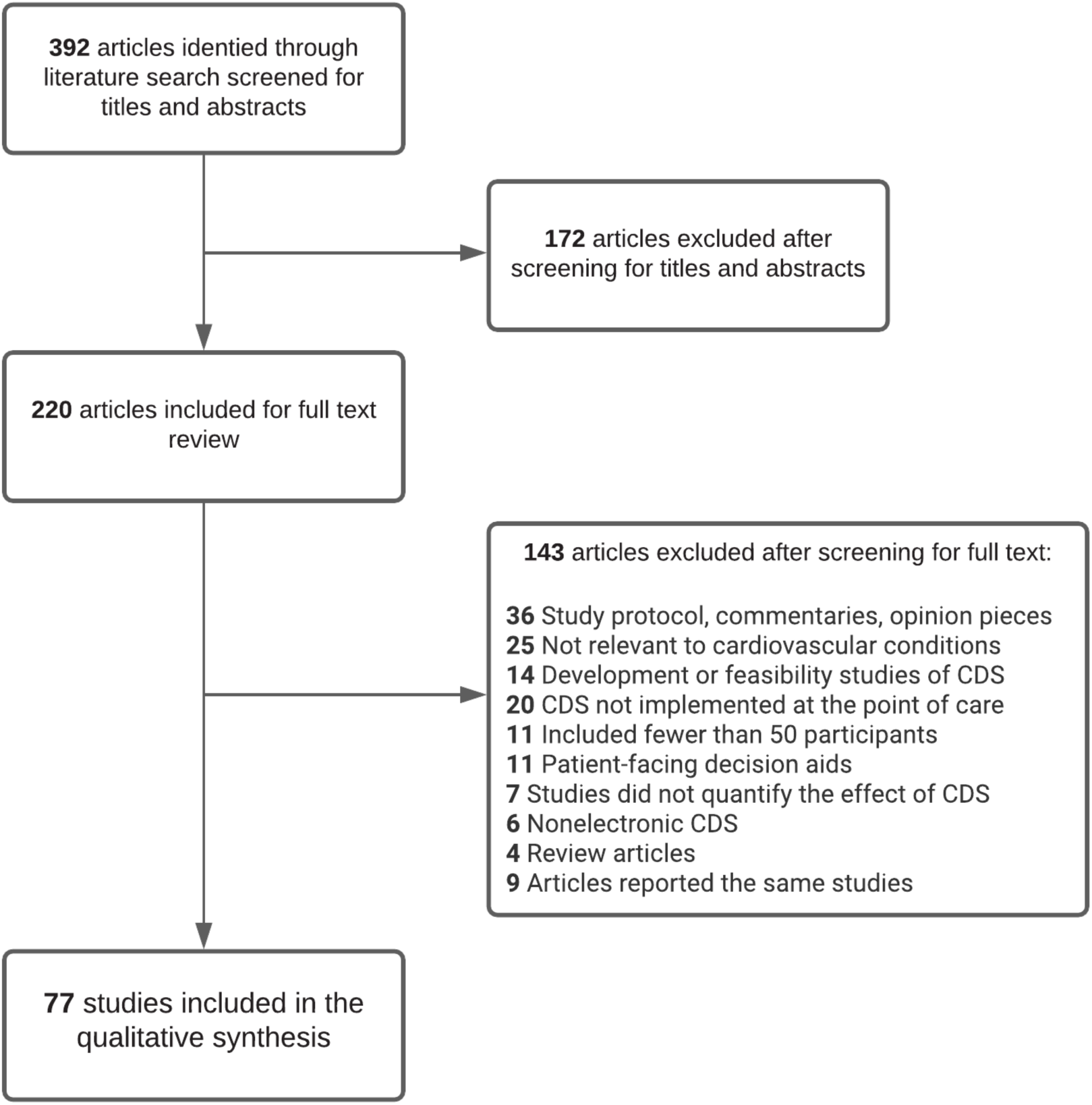
Flow of studies through the review process.

### Effect of the CDS on health and health delivery outcomes for CVD

As randomized design is considered the most rigorous method for evaluating CDS interventions,^24 25^ we focused on 58 RCTs to evaluate the effect of CDS and barriers to implementation.

#### Health care process outcomes

Overall, 45 of the 58 included RCTs assessed health care process outcomes for CVD (**Table 2 and eTable 2 in the Supplement**). Of these 45 studies, 23 reported a positive effect on improving the health care process outcomes of interest, 3 reported mixed effects, and 19 reported no significant effect. The vast majority (39 of the 45 studies) used usual care or no CDS as the control group without using an active comparator, while 3 studies compared against the same CDS with additional features and 3 studies compared CDS with other implementation strategies. Among studies that reported a positive effect, common process outcomes measured included CDS-recommended preventive service ordered or completion, clinical tests ordered or completion, and treatment prescribed. For example, a multi-center RCT of lipid management that enrolled 105 providers and 64,150 patients from 12 primary care clinics in the US found that CDS with guideline-based alerts improved the proportion of participants who were tested for hyperlipidemia by about 5%.^26^ Another large RCT of 197 Italian general practitioners and 21,230 patients found that the CDS intervention significantly increased the proportion of patients with diabetes prescribed antiplatelet drugs or lipid lowering drugs compared with usual care.^27^ A trial of pulmonary embolism diagnosis that enrolled 1,786 patients from 20 emergency departments in France found that the CDS intervention increased the proportion of patients who received an appropriate diagnostic work-ups by nearly 20%.^28^ Among studies that reported no significant effect of the CDS being studied on process outcomes, the most common factors identified contributing to the absence of effect were low use rate of CDS among providers, high baseline rates for the process measure of interest resulting in little room for improvement, improvement of outcomes in the control group because of taking part in the study (i.e., Hawthorne effect), and short follow-up periods to demonstrate the effect on outcomes.

**Table 2.** Summary of effect of CDS by outcome.

| Outcome | Number of studies that measured outcome | Summary of findings |
| --- | --- | --- |
| **Health care process outcomes (N=45)** |  |  |
| CDS-recommended preventive service ordered or completion, clinical tests ordered or completion, and treatment prescribed | 45 | 23 studies reported a positive effect on improving the health care process outcomes of interest, 3 reported mixed effects, and 19 reported no significant effect. |
| **User experience and other implementation outcomes (N=24)** |  |  |
| Provider acceptance and satisfaction | 13 | 8 studies reported good user experience, 3 reported neutral or unsatisfactory user experience, and 2 did not reported user experience results |
| Provider workload or efficiency | 0 | Evidence is lacking on the effect of CDS on provider workload or efficiency. |
| Patient knowledge | 1 | 1 study showed CDS intervention significantly improved provider knowledge in atrial fibrillation management. |
| Unintended consequence/<br>Adverse event | 12 | 9 studies reported harmful unintended consequences and 3 studies reported beneficial unintended consequences during CDS implementation. The most common unintended consequences reported were alert fatigue and distraction of current workflow. |
| **Clinical outcomes (N=41)** |  |  |
| CVD risk factor | 25 | 7 studies reported a positive effect of CDS on reducing CVD risk factors, 3 reported mixed effects, and 15 reported no significant effect. |
| CVD event | 21 | 4 studies reported a positive effect of CDS on reducing CVD events, 2 reported mixed effects, and 15 reported no significant effect. |
| **Economic outcomes (N=6)** |  |  |
| Cost | 4 | 4 studies showed CDS interventions were associated with reduced treatment costs, total costs, or hospitalization expense compared with control groups. |
| Cost-effectiveness | 2 | 2 studies reported that CDS was cost-effective in management of hypertension and diabetes. |
| **Health disparity outcomes (N=3)** |  |  |
| Race/ethnic differences | 3 | 3 studies showed a positive effect of CDS interventions in reducing CVD risk factors in minority populations. |

#### Clinical outcomes

Forty-one out of the 58 RCTs assessed clinical outcomes of CVD. Of these 41 studies, 10 reported a positive effect of CDS on improving clinical outcomes for CVD, 4 reported mixed effects, and 27 reported no significant effect. Common clinical outcomes assessed in these studies included CVD risk factor control (e.g., blood pressure and LDL cholesterol control rates), cardiovascular events, adverse events (e.g., bleeding), hospitalizations and mortality related to CVD. Compared with health process outcomes, fewer studies showed a positive effect on clinical outcomes. Among the 10 studies that reported positive effects, 7 studies showed a significant reduction in cardiovascular risk factor levels, including blood pressure and LDL cholesterol, and 3 studies showed a significant reduction in CVD events, including cardiovascular hospitalizations, major adverse cardiac events, and pulmonary embolism.

#### User experience and other implementation outcomes

A total of 13 studies assessed provider acceptance and satisfaction of the CDS intervention being studied. Of these, 8 reported good user experience, 3 reported neutral or unsatisfactory user experience, and 2 did not reported user experience results. The definition of provider acceptance was not consistent across studies. For example, in a multicomponent atrial fibrillation management intervention of 209 patients in China, provider acceptability was defined as provider satisfaction of the CDS tool;^29^ whereas, in another RCT of 39 clinicians and 781 patients in the Netherlands, provider acceptability was defined as provider acceptance of the CDS recommendations.^30^ Only one study assessed clinician knowledge or improved confidence in managing patient care as the intervention’s outcome.^29^ No study assessed the effect of CDS on clinician workload or efficiency.

Compliance with use of CDS or CDS recommendations was reported or discussed in 24 studies, many of which showed a large proportion of alerts were ignored by clinicians across studies. For example, in a RCT of stroke prevention that enrolled 39 clinicians and 781 patients from 18 Dutch general practices, clinicians ignored 60% of CDS alerts because they did not agree with the CDS recommendations.^30^

#### Economic outcomes

A total of 6 studies assessed economic outcomes and showed that CDS interventions were associated with reduced treatment costs, total costs, or hospitalization expense compared with control groups. Topics addressed in these studies included secondary prevention of stroke and vascular disease, oral anticoagulation prescription, management of stroke, hypertension, and diabetes. Two studies reported that CDS was cost-effective. In a RCT of 1,628 patients from 16 primary health center clusters in India, a CDS for managing hypertension was shown as highly cost-effective in resource constrained primary care settings.^31^ In another RCT of 3,391 patients in 55 primary care practices throughout the Netherlands, a multicomponent intervention combining task delegation, CDS, and feedback had a high cost-effectiveness ratio for improving cardiovascular risk for Type 2 diabetic patients.^32^

#### Health disparity outcomes

Evidence is lacking on the effect of CDS on reducing health disparities as it related to CVD risk factors. Three of the 58 studies included in this review measured health disparities as an outcome and assessed the impact of CDS on reducing this disparity.^33–35^ In a RCT of 573 patients in 3 primary care clinics in Veterans Affairs Medical Center in the US, a nurse-administered hypertension management intervention combining CDS and behavior program significantly reduced racial differences in blood pressure.^33^ In another large RCT of 38,725 patients in 60 primary healthcare centers in Australia, a CDS for cardiovascular disease risk management in primary health care reduced the gaps in care among aboriginal populations.^34^ As CVD and its risk factors disproportionally affect racial and ethnic minorities, studies evaluating CDS interventions on health disparity outcomes are highly needed.

#### Features associated with CDS effectiveness

Four of the 58 included trials directly compared a given CDS with the same system with additional features (e.g., providing additional information on patient symptoms), but they did not show a beneficial effect of the additional features.^30 36–38^ While evidence is lacking from the studies included in the present review, several previous reviews have identified design features that are closely related to the success of CDS. A large systematic review by Kawamoto et al analyzing 70 RCTs of CDS interventions found four features, including automatic provision of CDS as part of workflow, providing CDS at the time and location of decision making, providing recommendations in addition to assessments, and using computer-based CDS, were independent predictors for improving clinical practice.^7^ Other large systematic reviews echoed these findings and identified additional features associated with positive CDS effects. Specifically, CDS interventions were found to be more likely to succeed when automatically prompting users,^39^ minimizing the need for manual input of patient data, increasing the specificity and sensitivity levels of CDS advice,^40^ providing advice for patients in addition to practitioners, and evaluated by their developers.^41^ A recent systematic review of 108 RCTs showed that CDS requiring acknowledgement and documentation of reasons for not following the recommendations was associated with about 5% larger effects than CDS without this feature.^6^ The ability to execute the desired action through the CDS, considering alert fatigue in designing or delivering CDS were also associated with larger effects, but the incremental changes for these features were relatively small (**Figure 2**).

**Figure 2.**
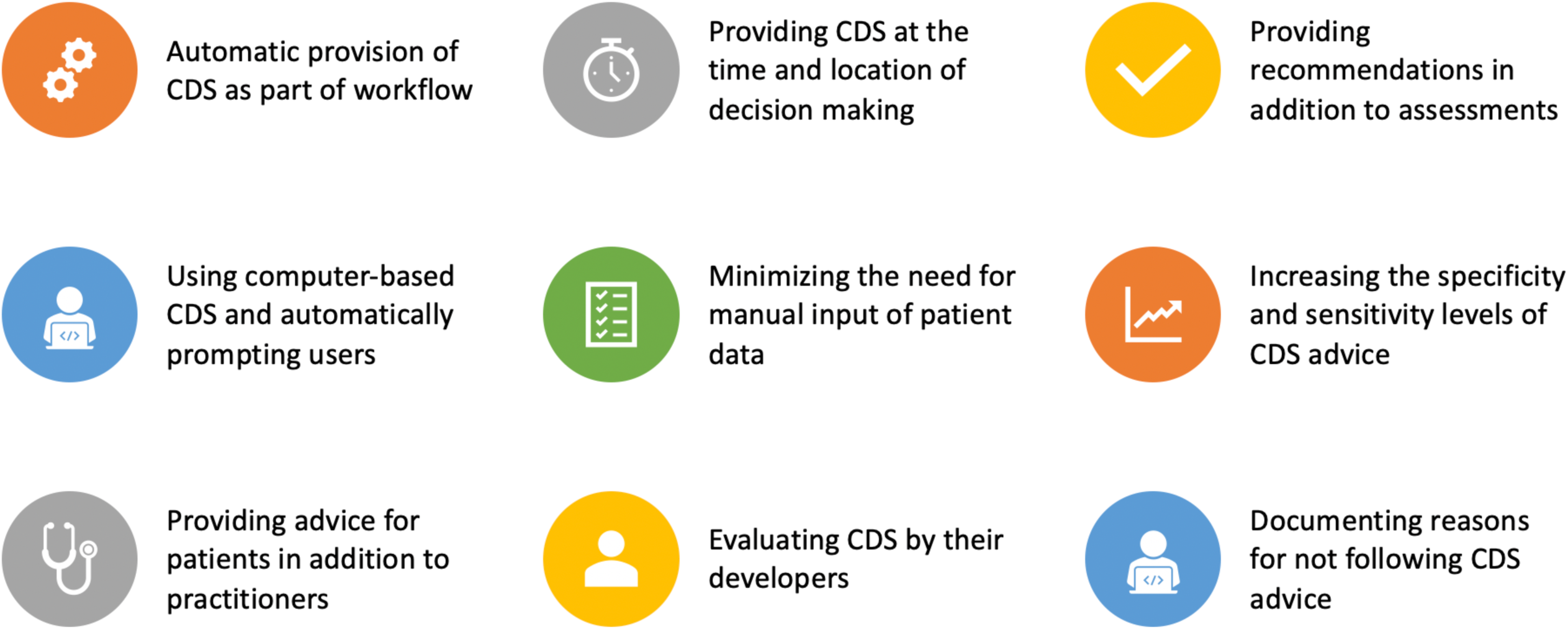
Clinical decision support features associated with larger effects on outcomes.

### Barriers to successful dissemination and implementation of CDS

Despite the great potential for CDS to improve care and patient outcomes in cardiovascular care, our review of the literature and prior systematic reviews consistently found low provider use of CDS across studies. The slow uptake of CDS in clinical practice highlights the need to understand barriers to successful dissemination and implementation of CDS in real-world settings.

From the 58 included RCTs, 31 reported barriers encountered during implementation of CDS interventions as assessed by post-intervention provider surveys or interviews. The most common barriers were time and resource constraints, lack of compatibility with workflow, alert fatigue, technical problems with CDS or EHR system, discordance between local guidelines and CDS recommendations, lack of trust in CDS recommendations, and complexity of real-world clinical management of CVD (**Table 3**). Notably, most of these barriers pointed to an underappreciation of the complex sociotechnical environment of real-world clinical settings in which the CDS was implemented.^42^ Other types of barriers mentioned in the studies were provider fear of doing harm, lack of provider awareness and training, lack of financial incentive, and lack of provider buy-in. As a result of these barriers, many CDS interventions failed to deliver the Five Rights of CDS (the right information to the right people in the right intervention formats through the right channels at the right points in workflow)^43^ and thereby limiting their impact to improve patient care.

**Table 3.** Common barriers encountered during CDS implementation and strategies to address barriers.

| Common barriers encountered during CDS implementation | Number of studies reported (N=31) | Example |
| --- | --- | --- |
| Time and resource constraints, lack of compatibility with workflow | 12 | In an RCT of hypertensive disorders in pregnancy that enrolled 2,286 patients in 16 Dutch hospitals, clinicians reported that the use of CDS was time-consuming and not well integrated into daily practice routines.<sup>44</sup> Ultimately, the CDS was used in only a quarter of eligible patients. |
| Alert fatigue, distraction from current workflow | 7 | In a large RCT of pharmacological management in high-risk cardiovascular patients that enrolled 197 general practitioners and 21,230 patients in Italy, it was reported that the main reason for discontinuation of the CDS use was due to a large number of alerts.<sup>27</sup> |
| Discordance between local guidelines and CDS recommendations, lack of trust in CDS recommendations | 6 | In the IMPART trial of venous thromboprophylaxis in acutely ill medical patients that enrolled 1,085 patients from 10 hospitals in Switzerland, CDS was used by only 30% of physicians; even when the CDS was used, only 75% of physicians followed the recommendation made by the tool.<sup>51</sup> The major reasons reported were provider distrust of CDS recommendations and discordance between local policies of thromboprophylaxis and the recommendations. |
| Complexity of real-world clinical management of CVD | 4 | A study of 1,493 patients from 15 primary care practices in the United Kingdom assessed the effect of an atrial fibrillation CDS tool in primary care. Post-visit survey results found that more than half of the providers discussed anticoagulation treatment with their patients, yet only 6% of them actually made a change in therapy at that visit given the nuance of real-world clinical situations.<sup>50</sup> |
| Technical problems with CDS or EHR-related issues | 5 | In a large RCT of a multifaceted intervention to prevent venous thromboembolism that enrolled 15,351 patients from 27 hospitals in France, the authors were only able to implement plugin reminders in two centers. Two centers had no EHR system, four had an EHR system incompatible with the plugin, five had hospital policies that did not allow plugin implementation.<sup>52</sup> |
| Others (provider fear of doing harm, lack of provider awareness and training, lack of financial incentive, lack of provider buy-in) | 7 | In a large RCT of stroke secondary prevention that enrolled 11,391 patients from 106 family practices from the United Kingdom, low utilization of CDS was reported and a qualitative process evaluation suggested physicians lack of awareness of the intervention.<sup>82</sup> |

| Common strategies to address barriers | Number of studies reported (N=23) | Example |
| --- | --- | --- |
| Integrating CDS into clinical workflow and the EHR | 13 | In a large RCT of hypertension management in adolescents that enrolled 31,579 patients from 20 primary care clinics within a large midwestern US health system, the CDS tool was designed to integrate with clinical workflow.<sup>58</sup> Provider alerts were only delivered when $\geq 2$ blood pressures were recorded at a visit and the average was $\geq 95$th percentile for age and sex based on the clinical guideline for hypertension management in children and adolescents. Audit and feedback to encourage nurses to repeat blood pressure measurements and providers to open the CDS along with in-person training sessions were conducted at intervention sites. As a result, a majority of providers using the CDS reported that it was useful for shared decision making. |
| Audit and feedback | 5 |  |
| Using more precise triggers to reduce alert fatigue | 4 |  |
| Obtaining provider buy-in and support from local leaders, conducting local consensus meetings, | 3 | In a RCT of 1,146 patients from 10 primary health clinics in India and Pakistan, a multi-component quality improvement intervention involving an EHR-based CDS and non-physician care coordinator has been showed to effectively improve achievement of diabetes care goals.<sup>83</sup> The authors mentioned that getting a provider buy-in at pre-adoption stage and showing evidential CDS benefit encouraged provider adoption of CDS. |
| Conducting ongoing provider training | 3 | In a RCT of 1,628 patients from 16 primary health center clusters in India, a CDS for managing hypertension was shown as highly cost-effective in resource constrained primary care settings.<sup>31</sup> Strategies used to facilitate CDS implementation included integrating CDS into existing clinical work flow, conducting provider training to ensure rigorous adherence of all sequential steps in CDS, engaging stakeholders in the CDS development process and using multicomponent intervention with concurrent advice to physicians and patients in institutional settings. |
| Others (Providing financial incentives, engaging front-line providers in the design of CDS, culturally adapting the CDS to study population) | 3 |  |

First, 12 studies reported time and resource constraints and lack of compatibility with workflow as barriers to CDS implementation. In an RCT of hypertensive disorders in pregnancy that enrolled 2,286 patients in 16 Dutch hospitals, clinicians reported that the use of CDS was time-consuming and not well integrated into daily practice routines.^44^ Ultimately, the CDS was used in only a quarter of eligible patients. Likewise, in a RCT of asthma and angina management that included 8,365 patients from 60 general practices in northeast England, use of CDS was low due to the challenge of integrating the CDS into clinical encounters where busy practitioners manage patients with multiple, complex conditions.^45^ In another large RCT of an intervention combining a point-of-care device for testing lipids and HbA1c with a web-based CDS for CVD risk assessment, 13,638 patients from 20 general practices in New Zealand were enrolled. Nurses reported feeling so time-pressured with their workload that having to wait for the test results and the sequential analysis of CVD risk by the machine was not good use of their time.^46^

Second, 7 studies reported alert fatigue (clinicians’ tendency to ignore repeated alerts) as a major negative unintended consequence when implementing CDS in real-world settings. This low signal-to-noise ratio for alerts was likely due to violations of one or more of the CDS Five Rights mentioned above, particularly not providing the right information at the right time in workflow. For example, in a large RCT of pharmacological management in high-risk cardiovascular patients that enrolled 197 general practitioners and 21,230 patients in Italy, it was reported that the main reason for discontinuation of the CDS use was due to a large number of alerts.^27^ In the CDS-AF study, 14,134 patients from 43 primary care clinics in Sweden were enrolled to study the effect of CDS on improving adherence to guidelines for anticoagulant therapy in patients with atrial fibrillation. Primary care physicians ignored the recommendation or made a decision that the patient would not benefit from therapy given the false alerts for patients who were at risk of stroke without appropriate treatment and additional workload imposed by the CDS tool.^47^ In another RCT of intraoperative hypotension that included 3,156 patients in the Hillcrest Hospital in Cleveland USA, anesthesiologists reported that they ignored CDS because the alert did not provide actionable recommendations (e.g., anesthesiologists were already aware of hypotension and doing their best to treat it) or the responses were ineffectual even the alerts provided additional information (e.g. anesthesiologists were distracted by the tool). Besides alert fatigue, another unintended consequence reported in the studies was distraction from current workflow, inappropriate alerts add to cognitive load which can lead to reduced provider efficiency and increased number of medical errors.^48 49^

Third, 10 studies reported discordance between local guidelines and CDS recommendations, lack of trust in CDS recommendations, and complexity of real-world clinical management of CVD as barriers to CDS implementation. A study of 1,493 patients from 15 primary care practices in the United Kingdom assessed the effect of an atrial fibrillation CDS tool in primary care. Post-visit survey results found that more than half of the providers discussed anticoagulation treatment with their patients, yet only 6% of them actually made a change in therapy at that visit given the nuance of real-world clinical situations.^50^ The most frequent documented explanations were patient preferences, anticoagulation therapy managed by another specialist, and concerns about increased fall risk in elderly patients. In the IMPART trial of venous thromboprophylaxis in acutely ill medical patients that enrolled 1,085 patients from 10 hospitals in Switzerland, CDS was used by only 30% of physicians; even when the CDS was used, only 75% of physicians followed the recommendation made by the tool.^51^ The major reasons reported were provider distrust of CDS recommendations and discordance between local policies of thromboprophylaxis and the recommendations.

Fourth, 5 studies reported technical problems with CDS or EHR-related issues as a barrier to CDS use. In a large RCT of a multifaceted intervention to prevent venous thromboembolism that enrolled 15,351 patients from 27 hospitals in France, the authors were only able to implement plugin reminders in two centers. Two centers had no EHR system, four had an EHR system incompatible with the plugin, five had hospital policies that did not allow plugin implementation.^52^ In another study of cardiovascular risk reduction that included 7,914 patients from 12 primary care clinics in Minnesota USA, implementation of CDS was delayed because the CDS containing the URL to display the CDS tool took longer to develop than anticipated.^53^

### Strategies to promote successful implementation

Despite multiple national efforts to advance CDS adoption and implementation,^54–57^ 35 out of the 58 RCTs included in this review did not explicitly mention adoption of any specific implementation strategies. Among the 23 studies that explicitly mentioned specific implementation strategies, the most common strategies were integrating CDS into clinical workflow and the EHR, audit and feedback, using more precise triggers to reduce alert fatigue, and conducting ongoing provider training (**Table 3**). Other strategies included obtaining support from local leaders, conducting local consensus meetings, providing financial incentives, engaging front-line providers in the design of CDS, and culturally adapting the CDS intervention to study population. Many of these strategies pointed to more deliberate processes for stakeholder engagement, buy-in, and continuous usability testing, which are endorsed by multiple professional organizations and federal agencies.^54–57^

Notably, successful studies that reported positive effectiveness of CDS and good user experience have applied specific strategies to optimize CDS dissemination and implementation. In a RCT of 7,914 patients from 12 primary care clinics in Minnesota USA, strategies used to achieve high CDS use rate included training of primary care physicians (PCP), engaging PCP and nursing leaders for workflow integration, monthly feedback at the clinic and PCP level of CDS use rates to intervention clinic managers and PCPs, and providing compensation twice to each intervention clinic nursing pool for clinics that sustained CDS use rates >75% of targeted patient.^53^ The trial showed significantly better annual rates of change in absolute 10-year cardiovascular risk in CDS clinics compare to clinics in the usual care arm of the trial and reported high provider satisfaction. In another trial of hypertension management in adolescents that enrolled 31,579 patients from 20 primary care clinics within a large midwestern US health system, the CDS tool was designed to integrate with clinical workflow.^58^ Provider alerts were only delivered when ≥2 blood pressures were recorded at a visit and the average was ≥95th percentile for age and sex based on the clinical guideline for hypertension management in children and adolescents.^59^ Audit and feedback to encourage nurses to repeat blood pressure measurements and providers to open the CDS along with in-person training sessions were conducted at intervention sites. As a result, a majority of providers using the CDS reported that it was useful for shared decision making. In the TORPEDO trial of 38,725 patients in 60 primary healthcare centers in Australia, strategies used to facilitate implementation included work flow integration, alignment with usual decision-making processes in the patient consultation, provision of treatment recommendations rather than just assessments, and repeated audit and feedback with explicit recommendations.^34^

### Current legal and regulatory environment for CDS

To advance toward widespread adoption and implementation of CDS, it is important to recognize and manage external factors, such as the policy, legal, and governance factors that affect the process of developing, disseminating, and implementing CDS interventions. An appropriate regulatory framework for CDS should seek to achieve an optimal balance between promoting technology innovation and protecting patients. To achieve this goal, the EU and the US use different approaches.^60^ In this section, we outlined the legal and regulatory landscape for CDS in the US and EU and described developments that are currently taking place.

#### The US approach

The current legal and regulatory landscape for CDS in the US is largely affected by the FDA medical device regulations. According to the 2019 FDA draft guidance on CDS,^61^ if a health care professional can independently review the basis for a recommendation, a CDS is not considered a medical device and hence not subject to FDA oversight. Specifically, the FDA used criteria from the 21st Century Cures Act^62^ and defined a Non-Device CDS as software that meets all four of the following criteria: (1) not intended to acquire, process, or analyze a medical image or a signal from an in vitro diagnostic device or a signal acquisition system; (2) intended for the purpose of displaying, analyzing, or printing medical information about a patient; (3) intended for the purpose of supporting or providing recommendations to a health care professional about prevention, diagnosis, or treatment of a disease or condition; (4) intended for enabling a health care professional to independently review the software recommendations to make a clinical diagnosis or treatment decision for patients. Device CDS, also known as Software as Medical Device (SaMD),^63^ are CDS that do not meet one or more of the four criteria above. The FDA applies a risk-based approach to provide oversight on SaMD as described below.

#### The FDA’s current risk-based approach for SaMD oversight

In 2017, the FDA adopted the International Medical Device Regulators Forum (IMDRF) framework in its risk based approach to SaMD regulation.^64^ This framework characterizes risk of SaMD based on two main factors: (1) the significance of information provided by the SaMD to the health care decision; and (2) the state of the health care situation or condition. As shown in **Table 4**, the categorization of risk occurs across a continuum, ranging from the lowest-risk software functions of informing clinical management for non-serious conditions (i.e., “inform x non-serious”) to the highest-risk software functions of diagnosing or treating critical conditions (i.e., “diagnose/treat x critical”).

**Table 4.**
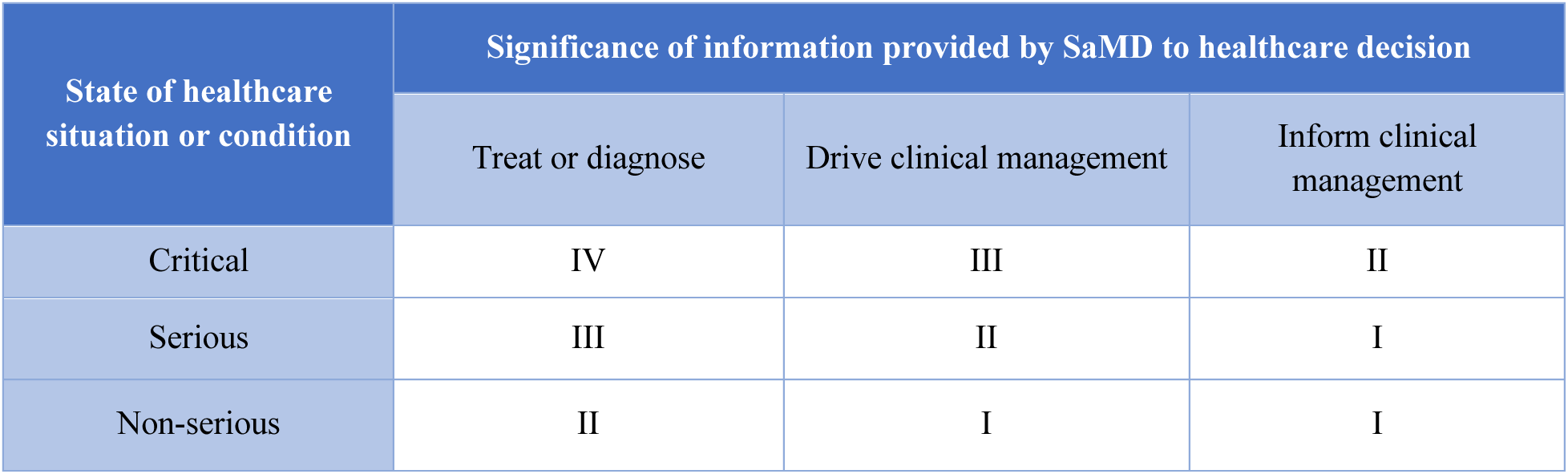
IMDRF framework for SaMD Risk Categorization.

According to the risk of SaMD, the FDA classifies devices into three distinct classes (Class I low-risk devices, Class II moderate-risk devices, and Class III high-risk devices) and regulates them accordingly (**Table 5**). The higher the risk, the stricter the control. Most of the Class I devices and all Class II devices are regulated by the FDA through the Pre-Market Notification (PMN) pathway (commonly known as 510(k) application). This is an expedited pathway that allows for devices to obtain marketing authorization if the sponsor can demonstrate substantial equivalence to an existing legally marketed device. Class III devices are devices involving the greatest risk and, therefore, are regulated by FDA through the more stringent Pre-Market Approval (PMA) pathway. This pathway requires extensive scientific evidence including technical, non-clinical laboratory, and clinical investigations to demonstrate a device’s safety and effectiveness prior to FDA approval. The FDA also has another pathway, De Novo pre-market review, for new low-to-moderate-risk devices without an equivalent predicate device. The De Novo pathway provides an opportunity to reclassify a novel Class III device to a Class I or Class II device, which then are subject to less stringent regulation. Specific examples of CDS in cardiovascular care for each device class are presented in **Table 5**.

**Table 5.** Summary of regulatory policy for CDS for which the intended user is health care professional.

| Can the user independently review the basis? | Is the CDS a medical device? | Class of medical device/level of risk |  | FDA regulation | Example in cardiovascular medicine |
| --- | --- | --- | --- | --- | --- |
| Yes | No | N/A |  | Not subject to FDA oversight | Software that uses a patient's diagnosis to provide health care professionals with current practice treatment guidelines for hypertension |
| No | Yes | Class I low-risk device | Software functions of "Inform x Non-Serious" | Enforcement Discretion | A medication reminder that provides alerts to healthcare providers for pre-determined medication dosing schedules (Product Code NXQ) |
|  |  |  | Others | Pre-market notification (PMN or 510k) | A machine learning algorithm, for which the logic and inputs are not explained, that identifies hospitalized diabetic patients at increased risk of postoperative cardiovascular events |
|  |  | Class II medium-risk device |  | Pre-market notification (PMN or 510k) | Software that processes and analyzes imaging data to identify the presence of certain CVD |
|  |  | New device belongs to Class I or Class II |  | De Novo | A screening device that analyzes images and directly refers patients to cardiologists |
|  |  | Class III high-risk device |  | Pre-market approval (PMA) | An implantable pacemaker for cardiac patients |

#### FDA’s new Software Precertification Program

There is concern that the traditional device approval/clearance pathway discussed above is not suited to the faster, more iterative design and development cycles of devices, which takes weeks-to-months rather than the months-to-years long cycle of more traditional medical products. As such, the FDA released the Digital Health Innovation Action Plan in July 2017,^65^ announcing that it was reimagining the approach to digital health medical devices. In 2018, the FDA started the Software Precertification Pilot Program^66^ which is considered a voluntary alternative to the traditional PMN/PMA pathway to provide more streamlined oversight of digital devices and accelerate their time to market. This program aims to shift the focus from solely pre-market evaluation to evaluating both the product and the company by continuously monitoring the real-world performance of its products once they have reached the market. Although the program is still in the development phase, the current version has 4 interdependent components:^67^ (1) excellence appraisal and precertification, (2) review pathway determination, (3) streamlined pre-market review process, and (4) real-world performance (**Table 6**).

**Table 6.**
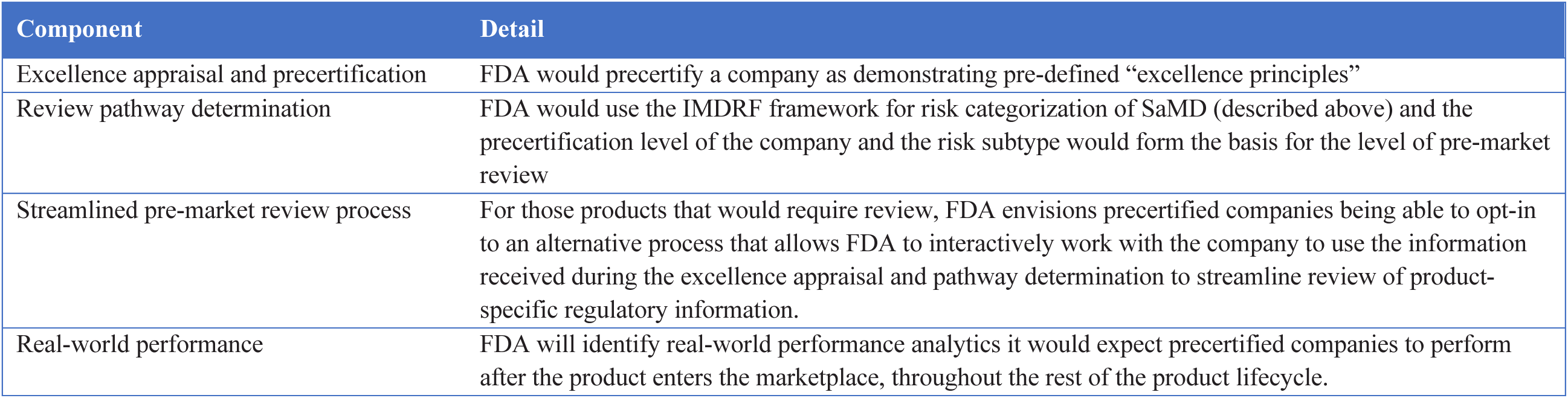
Four interdependent components of FDA’s Software Precertification Program.

To respond to the COVID-19 pandemic and to provide more efficient oversight in the midst of the pandemic, the FDA has also suspended requirements for certain ‘lower risk device’ software. For the subset of ‘higher-risk’ digital health devices, the FDA is using the Emergency Use Authorization process to help expand access to medical products for use during the pandemic.

#### The EU approach

The EU is reforming the legal framework for medical devices with a number of new legislative reforms (General Data Protection Regulation [GDPR], Cybersecurity Directive, and Medical Devices Regulation). While reform is a gradual process, the GDPR and the Cybersecurity Directive enacted in May 2018 have already begun to have an impact.

The definition of medical device in the EU includes any kind of software, intended by the manufacturer to be used for human beings for the purpose, among others, of diagnosis, prevention, monitoring, treatment, or alleviation of disease.^68^ This definition has been endorsed by the MEDDEV guidelines drafted by the European Commission to guide stakeholders in complying with legislation related to medical devices.^69^

The regulatory framework for CDS in the EU is largely affected by three directives on medical devices created in the 1990s.^68 70 71^ These directives require manufacturers to comply with a number of essential requirements depending on the risk classification of the device and to ensure that the produced devices are fit for their intended purpose. Whether the essential requirements have been met can be assessed either by the manufacturer or by an independent accredited certification organization appointed by the competent authorities of EU Member States.

Recognizing that the existing directives do not fit with new, evolving technologies, the EU issued a new Medical Devices Regulation in May 2017.^72^ This new regulation, officially applied from May 2020, extends the scope to include a wider range of products, extends the liability in relation to defective products, strengthens requirements for clinical data and traceability of the devices, provides more rigorous monitoring of independent certification organizations, and improves transparency through making information relating to medical devices available to the public. Different from directives that require national legislation to implement their purposes, the new regulation is applied directly in EU Member States without the need for national legislation to implement.

#### Data privacy considerations for CDS using patient data

Developers and implementers of CDS using patient data should be aware of the various laws that protect health information privacy.^73^ In the US, the Health Insurance Portability and Accountability Act of 1996 (HIPAA) established national standards regarding the protection of patient health information.^74^ Development of CDS using data from patients that have consented to its use is permitted, and HIPPA requires that patients be given the right to direct any covered entity to transmit a copy of their medical records to a designated entity of the individual’s choice.^75^ However, there are several exceptions to the requirement of patient consent under HIPAA. For example, protected health information may be used by CDS without patient consent to support selected quality improvement activities, clinical guideline development, and population-based activities relating to reducing costs.^76^ Besides federal laws, several states in the US have their own data privacy protection laws and regulations that must be followed. Thus, companies that plan to operate in multiple states need to ensure their compliance with each state’s laws.

In the EU, the GDPR enables individuals to control their own health data. In particular, the GDPR has regulations that outline individuals have the “right to an explanation” when it comes to machine learning algorithms.^77 78^ For automated decision-making, individuals have the right to object to any decision made about them if that decision is based purely on automated processing.^79^ Individuals also have the right to obtain information about the logic involved in the automated decision-making system, its significance, and any resulting consequences. Diagnostic tools previously not considered medical devices may be considered medical devices under the new regulations if they have a purpose of “prediction and prognosis.”^80^

### Guidelines

High-quality CDS studies are needed to learn the best ways to apply CDS systems to achieve important improvements in healthcare delivery and outcomes, however limited guidance is available for designing and reporting CDS studies. Kawamoto et al recently published an article that summarized the key issues encountered in previous CDS studies and proposed 13 recommendations for research and reporting of CDS studies (**Table 7**).^81^ If adopted, these recommendations could help improve the quality of CDS studies and ultimately fulfill the CDS promise of more efficient and effective care. Eventually, CDS interventions, especially in high-stakes settings, should be evaluated with the same rigor as other therapeutics.

**Table 7.**
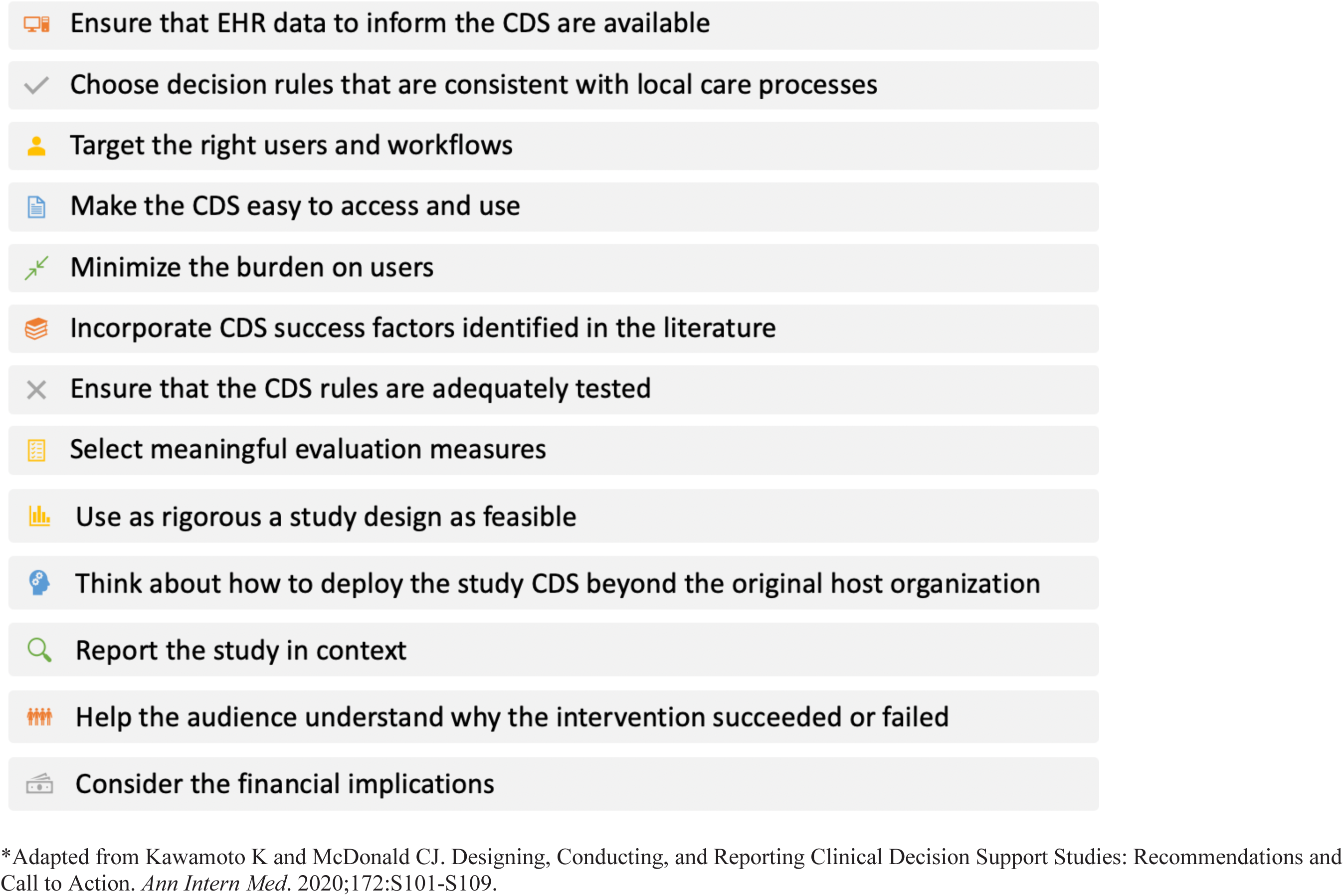
Recommendations for research and reporting of CDS studies.

In addition to reporting guidelines, guidance on best practices in dissemination and implementation of CDS based on real-world experience is also lacking. The 2017 National Academy of Medicine on Optimizing Strategies for Clinical Decision Support highlighted the importance of disseminating best practices for CDS.^57^ The report proposed convening expert groups to cultivate, plan, and direct the publication of actionable implementation guides that draw upon existing efforts to delineate best practices in implementation and platform integration approaches for different delivery systems; CDS management approaches for organizing multi-stakeholder CDS implementation and governance committees, and for clinicians and health systems of various sizes/resources; and usability recommendations for usable, practical, workflow-supportive CDS for various situations.

## Conclusion

Despite a rapid increase in publications of CDS studies in cardiovascular care over the past two decades, this review showed marked heterogeneity in the design, implementation, and evaluation of CDS interventions. Evidence demonstrates the positive effectiveness of some CDS in improving CVD care process outcomes, such as screening and preventive care services, ordering recommended clinical tests, and prescribing recommended treatments to mitigate the risk of CVD. The context in which the CDS is applied may strongly influence the results, highlighting the concomitant importance of implementation science. The results on improvement in CVD-related clinical outcomes are more mixed and evidence is lacking on the effectiveness of CDS on improving other implementation outcomes, economic outcomes, and health disparities as they relate to CVD. The uptake of CDS in CVD care remains slow and barriers to implementation exist at multiple levels. Many of these barriers are due to a lack of adequate understanding of the end users’ needs, a lack of EHR integration, and usability issues. Therefore, widespread adoption of CDS will require aggressively seeking a better understanding of what the right information is and when and how it should be delivered to the right person. Applying user-centered design to obtain deep understanding of those who use CDS and integrating their needs specific to the decision at hand into the CDS design with continuous user testing is important in achieving this goal.

Taken together, the available evidence shows that CDS has yet to realize its potential in cardiovascular care. Learning from more than two decades of evidence and establishing guidelines and best practices in designing, implementing, evaluating, and reporting CDS interventions are crucial to fulfill the promise of CDS to enhance care delivery, accelerate system-wide continuous learning, and improve health care outcomes for CVD.

## Data Availability

All data are included in the online supplemental material.

## Research questions

- What is the effectiveness of CDS on patient-centered outcomes, economic outcomes, and health disparity outcomes related to cardiovascular diseases? Can cardiovascular CDS achieve long-term sustainability?
- Can CDS be effective for other members of the healthcare team (such as nurses and pharmacists)?
- How can CDS for cardiovascular diseases be expanded to accommodate multiple comorbid conditions simultaneously and how can new CDS products be integrated with existing electronic health record systems?
- Considering implementation of CDS for cardiovascular diseases in real-world settings ranging from small physician practices to large health systems and across a variety of workflows, which implementation models are more efficient than others and in what settings?

## Patient involvement

No patients were asked for input in the creation of this article.

## Contributors

YL, ERM, and HMK conceived and designed this review. YL conducted the search and selected the studies for inclusion. YL drafted the manuscript, and ERM and HMK edited and approved the final version.

