## Supplemental material for "Clinical Decision Support in Cardiovascular Medicine: Effectiveness, Implementation Barriers, and Regulation"

**Online Supplement**

### Table of Contents

eTable 1. Characteristics of 77 included studies ........ 2

eTable 2. Effectiveness of CDS, barriers to implementation, and strategies to improve implementation in 58 randomized trials ........ 9

### eTable 1. Characteristics of 77 included studies

| Study ID | Country in which the study conducted | Study design | Duration of study (months) | Study setting | Sample size | Clinical domain of intervention | Intervention group | Control group |
|---|---|---|---|---|---|---|---|---|
| Dregan 2014[1] | United Kingdom | Cluster RCT | 12 | 106 Clinical Practice Research Datalink family practices in UK | 106 clinics, 11391 patients | Preventive care of CVD | CDS with guideline-based reminders and recommendations in EHRs for secondary prevention of stroke and vascular disease | Usual care |
| Mazzaglia 2016[2] | Italy | Cluster RCT | 12 | 197 Italian general practitioners | 197 Providers, 21230 patients | CVD management | CDS with guideline-based alerts for management of high-risk CVD patients | Usual care |
| Lester 2006[3] | United States | RCT but randomized at the individual level | 12 | 14 primary care physicians in an academically affiliated practice with an electronic health record | 235 patients, 14 clinics | Preventive care of CVD | CDS with guideline-based email reminders to doctors for hypertensive patients | Usual care |
| Montgomery 2000[4] | United Kingdom | Cluster RCT | 24 | 27 general practices in Avon, England | 27 providers, 614 patients | Screening of CVD risk factors | CDS incorporated with EHR for risk assessment of hypertensive patients plus risk chart assessment | Usual care, chart assessment only |
| Panjasawatwong 2015[5] | United States | RCT but randomized at the individual level | 12 | Hillcrest Hospital in Cleveland, USA | 3156 patients | Preventive care of CVD | CDS with alert for intraoperative hypotension | Usual care |
| Karlsson 2018[6] | Sweden | Cluster RCT | 12 | 43 primary care clinics in Sweden | 43 clinics, 14134 patients | Preventive care of CVD | CDS with alerts to physicians for patients with AF at risk of stroke without appropriate treatment | Usual care |
| Eckman 2016[7] | United Kingdom | Cluster RCT | 10 | 15 primary care practices in UK | 15 clinics, 1493 patients | Preventive care of CVD | CDS with guideline-based alerts for use of thromboprophylaxis in AF patients | Usual care |
| Bucher 2010[8] | Switzerland | Cluster RCT | 18 | 7 centers that specialize in the care of HIV-infected individuals in Switzerland | 7 centers, 165 providers, 3266 patients | Preventive care of CVD | CDS with guidelines for CHD and risk profile of patients infected with HIV | CDS with guidelines for CHD alone |
| Weir 2003[9] | Scotland | Cluster RCT | 6 | 16 hospitals in Scotland | 16 hospitals, 1952 patients | Preventive care of CVD | CDS with EHR based risk assessment and management guidelines | Usual care |
| Keeffe 2005[10] | United States | Cluster RCT | NR | 2 VA medical centers in the US | 2 VA centers, 52 providers | CVD management | CDS with alerts for guideline-based recommendations and patient symptom data | CDS without patient symptom data |
| Gill 2009[11] | United States | Cluster RCT | 12 | 12 primary care clinics in US | 12 clinics, 105 providers, 64150 patients | Preventive care of CVD | CDS with guideline-based alerts in EHRs for lipid management in primary care setting. | Usual care |
| Beeler 2014[12] | Switzerland | Cluster RCT | 2 | A university hospital in Switzerland | 15736 patients | Preventive care of CVD | CDS with alerts for initiation of prophylaxis order in patients who did not receive an order in 6hr of admission or transfer | Usual care |
| Luitjes 2018[13] | Netherlands | Cluster RCT | 13 | 16 Dutch hospitals | 2286 patients, 16 clinics | CVD management | CDS with guideline-based recommendations along with audit and feedback | Professional audit and feedback |
| Frances 2001[14] | United States | Cluster RCT | 3 | A Veterans Affairs Medical Center in San Francisco, USA | 1 center, 66 providers, 730 patients | Preventive care of CVD | CDS with reminders for patients with coronary artery disease | Usual care |

| Study ID | Country in which the study conducted | Study design | Duration of study (months) | Study setting | Sample size | Clinical domain of intervention | Intervention group | Control group |
|---|---|---|---|---|---|---|---|---|
| Hetlevik 1999[15] | Norway | Cluster RCT | 18 | General practice in Sør- and Nord-Trøndelag counties in Norway | 2239 patients, 55 providers | Screening of CVD risk factors | CDS with guideline-based assistance in diagnostics, history and treatments for hypertensive patients | Usual care |
| Kharbanda 2018[16] | United States | Cluster RCT | 24 | 20 primary care clinics within a large Midwestern health system | 20 clinics, 31579 patients | Both CVD diagnosis and CVD management | CDS screening for high blood pressure in primary care setting and CDS with guideline-based intervention | Usual care |
| O'Connor 2014[17] | United States | Cluster RCT | 14 | Kaiser Permanente Colorado and HealthPartners Medical Group in Minnesota | 32 providers, 19960 patients | Preventive care of CVD | Two intervention arms including (1) PPL-EMR intervention in which cases were assigned to each physician based on observed patterns of care in the EHR in the previous year, or (2) PPL-ASSESS intervention in which cases were assigned to each physician based on their performance on four standardized assessment cases | Usual care |
| Ranta 2015[18] | New Zealand | Cluster RCT | 15 | 56 Primary care clinics in New Zealand | 291 patients, 56 clinics | CVD management | CDS with EHR based guided management of TIA/stroke | Usual care |
| Nendaz 2010[19] | Switzerland | Cluster RCT | 4 | 10 hospitals in Switzerland | 10 hospitals, 1085 patients | Preventive care of CVD | Two intervention arms, including (1) CDS with guideline-based alerts to start thromboprophylaxis and (2) pocket cards | Usual care |
| Sperl-Hillen 2018[20] | United States | Cluster RCT | 14 | 12 primary care clinics in Minnesota, US | 12 clinics, 7914 patients | Preventive care of CVD | CDS with guideline based EHR reminders for treatment of CVD risk factors | Usual care |
| Chaturvedi 2019[21] | United States | Cluster RCT | 15 | 3 stroke centers in the US | 3 centers 309 patients | Preventive care of CVD | CDS with electronic alerts for OAC medication prescription for patients with AF by calculating CHA2DS2-VASc score | Usual care |
| Roy 2016[22] | France | Cluster RCT | 3 | 27 community and academic hospitals in France | 27 hospitals, 15351 patients | Preventive care of CVD | Multicomponent intervention consisting of CDS with guideline-based alerts for VTE prevention along with doctor education and pocket cards | Usual care |
| Fitzmaurice 2000[23] | United Kingdom | Cluster RCT | 12 | 12 primary care practices in Birmingham, England | 376 patients | CVD management | Intervention package involving near-patient testing and CDS with guideline-based reminders for use of oral anticoagulation | Intrapractve controls (within the intervention cluster) and Inter-practice controls getting usual care |
| Holbrook 2011[24] | Canada | RCT but randomized at the individual level | 12 | 49 primary care practices in Ontario, Canada | 49 clinics, 1102 patients | Preventive care of CVD | CDS with guideline-based reminders for assessment of risk of CVD | Usual care |
| Gilutz 2009[25] | Israel | Cluster RCT | 47 | 112 clinics in the Clalit Health Services in Israel. | 112 clinics, 204 providers, 7448 patients | Preventive care of CVD | CDS with monitoring and treatment alerts for patients with CAD. | Usual care |

| Study ID | Country in which the study conducted | Study design | Duration of study (months) | Study setting | Sample size | Clinical domain of intervention | Intervention group | Control group |
|---|---|---|---|---|---|---|---|---|
| Arts 2017[26] | Netherlands | Cluster RCT | 11 | 18 Dutch general practices | 18 clinics, 39 providers, 781 patients | CVD diagnostics | CDS with guideline-based recommendations for atrial fibrillation that uses the CHA2DS2-VAsc for stroke risk stratification, with and without documenting justification for reasons for not following CDS recommendation | Usual care |
| vanEngen-Verheul 2015[27] | Netherlands | Cluster RCT | 17 | 18 cardiac rehabilitation clinics in the Netherlands | 18 clinics, 14847 patients | CVD management | Multicomponent intervention consisting of CDS with web-based audits and feedback, education outreach | Usual care |
| Hetlevik 2000[28] | Norway | Cluster RCT | 13 | 29 general practice in Norway | 29 clinics, 1034 patients, 53 providers | Preventive care of CVD | CDS with guideline-based alerts for care of patients with type 2 diabetes | Usual care |
| Wells 2017[29] | New Zealand | Cluster RCT | 8 | 20 general practices in New Zealand | 20 clinics, 13638 patients | Screening of CVD risk factors | CDS for CVD risk assessment | Usual care |
| Eccles 2002[30] | United Kingdom | Cluster RCT | 24 | 60 general practices in north east England | 60 clinics, 8365 patients | CVD management | CDS with guideline-based reminders for management of asthma and angina in adults in primary care | Usual care |
| Cheng 2018[31] | United States | RCT but randomized at the individual level | 31 | Four county hospitals in the Los Angeles County Department of Health Services public healthcare system | 404 patients | Preventive care of CVD | CDS issued guideline-based reminders to NPs and PAs for patients with a TIA or ischemic stroke in the last 90 days. | Usual care |
| vanDoorn 2018[32] | Netherlands | Cluster RCT | 19 | 38 general practices in the region of Utrecht, the Netherlands | 38 clinics, 2255 patients | Preventive care of CVD | CDS with guideline-based initiation of prophylaxis for stroke prevention in AF patients | Usual care |
| Subramanian 2004[33] | United States | Cluster RCT | 12 | 2 Veterans Affairs medical centers | 2 centers, 720 patients | CVD management | CDS with suggestions based on EHR data and symptom data obtained from patient questionnaires | CDS with EHR data alone |
| Anchala 2015[34] | India | Cluster RCT | 5 | 16 primary health center clusters from Telangana state, India | 16 clinics, 1628 patients | CVD management | CDS helping physicians to (1) evaluate risk factors for CVD (2) classify risk level (3) follow guideline-based drug management (4) give alerts on lifestyle changes and medication adherence for management of hypertensive patients. | Chart Based System with the guidelines and lifestyle advices printed as a poster format |
| Roy 2009[35] | France | Cluster RCT | 12 | 20 emergency departments in France | 20 emergency departments, 1786 patients | CVD diagnostics | CDS with diagnostic guidelines for the diagnosis of PE in ED | Poster and pocket cards with diagnostic strategies |
| Tian 2015[36] | China and India | Cluster RCT | 26 | 47 villages in China and India | 2086 patients | Screening of CVD risk factors | Multicomponent intervention consisting of CDS with guideline-based recommendations for medication use and lifestyle modification in people with high cardiovascular risk, patient education | Usual care |
| Guo 2017[37] | China | Cluster RCT | 4 | Chinese PLA General Hospital and Meishan City People's Hospital | 209 patients | Preventive care of CVD | Multicomponent intervention consisting of mobile based CDS with guideline-based reminders for patients with AF, patient education and adherence tracking | Usual care |

| Study ID | Country in which the study conducted | Study design | Duration of study (months) | Study setting | Sample size | Clinical domain of intervention | Intervention group | Control group |
|---|---|---|---|---|---|---|---|---|
| Goud 2009[38] | Netherlands | Cluster RCT | 6 | 21 cardiac rehabilitation centers in the Netherlands | 21 clinics, 2787 patients | CVD management | CDS with guideline-based reminders incorporated in EHR for management of cardiac rehabilitation patients | Usual care |
| B. Sussman 2018[39] | United States | Cluster RCT | 9 | A Veterans Affairs clinic in Michigan, USA | 43 providers, 3787 patients | CVD management | Multicomponent intervention consisting of a personalized decision support tool, an educational program, a performance measure, and an audit and feedback system for statin use in patients according to 2013 guidelines. | Usual care |
| Cleveringa 2008[40] | Netherlands | Cluster RCT | 29 | 55 primary care practices throughout the Netherlands | 55 clinics, 3391 patients | Screening of CVD risk factors | Multicomponent intervention consisting of 1) a diabetes consultation hour run by a practice nurse,2) a CDS containing a diagnostic and treatment algorithm based on the Dutch primary care of type 2 diabetes guidelines and providing patient-specific treatment advice, 3) a recall system, and 4) feedback at both practice and patient-level every 3 months regarding the percentage of patients meeting the treatment targets | Usual care |
| Peiris 2015[41] | Australia | Cluster RCT | 8 | 60 primary healthcare centers in Sydney Australia | 60 clinics, 38725 patients | CVD management | CDS with guideline-indicated risk factor measurements and guideline-indicated medications for those at high cardiovascular disease risk | Usual care |
| Peiris 2019[42] | India | Cluster RCT | 24 | 18 primary health center clusters in rural Andhra Pradesh, India | 18 clinics, 62254 patients | Preventive care of CVD | A multifaceted intervention to support PHC doctors in the provision of guidelines-based assessment and management of CVD risk. | Usual care |
| Ali 2016[43] | India and Pakistan | RCT but randomized at the individual level | 42 | 10 urban clinics, nine in India and one in Pakistan | 10 clinics, 1146 patients | CVD management | Multi-component quality improvement project: non-physician care coordinators (to motivate patients) and EHR-based CDS (recommended care prompts to physicians) to improve diabetes care | Usual care |
| Johnston 2019[44] | United States | RCT but randomized at the individual level | 76 | 63 hospitals in the US | 63 hospitals, 1151 patients | Preventive care of CVD | CDS to maintain blood glucose concentration of 80-130mg/dL with intravenous insulin infusion - intensive care | Usual care |
| Zhu 2018[45] | China | RCT but randomized at the individual level | 13 | An urban community health center in Guangzhou China | 134 patients | CVD management | CDS with guideline-based alerts used by nurses for management of hypertensive patients. | Usual care |
| McKinstry 2013[46] | United Kingdom | RCT but randomized at the individual level | 32 | 20 primary care practices in south east Scotland | 20 clinics, 401 patients | CVD management | CDS with alerts based on patient BP measurements with optional patient decision support | Usual care |
| Hicks 2008[47] | United States | RCT but randomized at the individual level | 19 | 8 community-based and 6 hospital-based primary care practices affiliated with a large urban academic medical center | 2027 patients | CVD management | CDS with guideline-based reminders for hypertensive patients | Usual care |

| Study ID | Country in which the study conducted | Study design | Duration of study (months) | Study setting | Sample size | Clinical domain of intervention | Intervention group | Control group |
|---|---|---|---|---|---|---|---|---|
| Brocklehurst 2018[48] | United Kingdom | RCT but randomized at the individual level | 43 | 24 maternity units in the UK and Ireland. | 24 clinics, 47602 patients | CVD diagnostics | CDS that assesses fetal heart signals and gives color coded alerts for assessment of pregnant women at >35 weeks' gestation | Usual care |
| Piazza 2009[49] | United States | RCT but randomized at the individual level | 16 | 25 medical centers throughout the United States | 2493 patients | Preventive care of CVD | CDS with guideline-based alerts for patients at high risk of VTE | Usual care |
| Kucher 2005[50] | United States | RCT but randomized at the individual level | 40 | Brigham and Women's Hospital in the USA | 2506 patients | Preventive care of CVD | CDS with guideline-based alerts for risk of deep vein thrombosis | Usual care |
| Bhavnani 2018[51] | India | RCT but randomized at the individual level | 14 | Sri Satya Sai Institute of Higher Medical Sciences in Bangalore, India | 253 patients | CVD diagnostics | mHealth diagnostic strategy including pocket-echocardiography, smartphone-connected-electrocardiogram blood pressure and oxygen measurements, activity monitoring, and portable brain natriuretic peptide laboratory testing at point-of-care | Usual care |
| Mahler 2015[52] | United States | RCT but randomized at the individual level | 17 | A tertiary care academic medical center in North Carolina, USA | 282 patients | CVD management | CDS with guideline-based reminders for testing and management of chest pain | Usual care |
| Tutino 2017[53] | China | RCT but randomized at the individual level | 20 | 6 tertiary hospitals in China | 3586 patients | Screening of CVD risk factors | Multicomponent intervention combining structured assessment, risk stratification, personalized reporting and decision support | JADE Group (Joint Asia Diabetes Evaluation) - module to facilitate structured follow-up visits, along with identical built-in protocols as the intervention group |
| Jackson 2012[54] | United States | RCT but randomized at the individual level | 38 | 3 primary care clinics in Veterans Affairs Medical Center in Durham, North Carolina. | 573 patients | CVD management | Three intervention arms including (1) nurse-administered, physician-directed medication management intervention, utilizing a validated CDS; (2) nurse-administered, behavioral management intervention; 3) combined behavioral management and medication management intervention | Usual care |
| Huebschmann 2012[55] | United States | RCT but randomized at the individual level | 11 | 7 clinics in University of Colorado Hospital primary care system | 7 clinics, 591 patients | CVD management | CDS with guideline-based reminders for hypertension management | Usual care |
| Bosworth 2007[56] | United States | RCT but randomized at the individual level | 18 | 3 primary care clinics in Veterans Affairs Medical Center in Durham, North Carolina. | 3 clinics, 600 patients | CVD management | Three intervention arms including (1) tailored behavioral intervention, (2) medication management intervention using a validated CDS, and (3) combined behavioral and medication management intervention | Usual care |

| Study ID | Country in which the study conducted | Study design | Duration of study (months) | Study setting | Sample size | Clinical domain of intervention | Intervention group | Control group |
|---|---|---|---|---|---|---|---|---|
| Smith 2008[57] | United States | RCT but randomized at the physician level | 29 | 6 primary care practices in Rochester, MN | 6 clinics, 97 providers, 639 patients | Screening of CVD risk factors | CDS with guideline-based alerts for risk factors of CVD | Usual care |
| Kline 2014[58] | United States | RCT randomized at physician level | 11 | 3 academic emergency departments and 1 community hospital | 3 academic emergency departments and 1 community hospital, 550 patients | CVD diagnostics | CDS with pre-test probability for both acute coronary syndrome and pulmonary embolism and suggested clinical actions designed to lower radiation exposure and cost. | Usual care |
| Vinson 2018[59] | United States | Controlled pragmatic trial | 15 | 21 community EDs of Kaiser Permanente Northern California, USA | 21 emergency departments | CVD management | CDS with guideline-based diagnosis and treatment suggestions | Usual care |
| Baroletti 2008[60] | United States | Cohort Study | 30 | Brigham and Women's Hospital, USA | 866 patients | Preventive care of CVD | CDS using electronic alerts for prevention of VTE | All endpoints in the present cohort study were compared to the alert arm from the original trial |
| Robson 2014[61] | United Kingdom | Cross-sectional | 24 | 143 general practices in London | 3964 patients in 2011 and 4168 patients in 2013 | Preventive care of CVD | CDS with guideline-based reminders for anticoagulation in AF patients | Usual care |
| Beltran 2017[62] | Spain | Pre-post study | 4 | 2 primary-care centers in Madrid, Spain | 2 primary care clinics, 162 patients | CVD management | CDS with guideline-based recommendations for patients with type 2 diabetes | No control group |
| LaBresh 2008[63] | United States | Pre-post study | 11 | 99 hospitals in Georgia, Massachusetts, Michigan, Ohio, Florida, Arizona, northern California, southeastern Pennsylvania, USA | 99 clinics, 18410 patients | CVD management | CDS with guideline-based reminders for use of thrombolytic medications and antithrombotic medications in TIA/Stroke | No control group |
| Bookman 2017[64] | United States | Pre-post study | 17 | 5 emergency departments in a healthcare system | 235858 patient visits, 5 emergency departments | CVD diagnostics | CDS with scoring tools embedded in EHR for guiding the use of CTs for patients with head injury, C-spine injury or PE | No control group |
| Drescher 2011[65] | United States | Pre-post study | 3 | A university-affiliated community hospital ED, USA | 404 patients | CVD diagnostics | CDS with guidelines for evaluation of PE in EDs for guiding use of CT angiography | No control group |
| Aase 1999[66] | Norway | Pre-post study | 5 | Emergency room at the Central Hospital of Akershus, Norway | 32 providers, 493 patients | CVD diagnostics | CDS used by the emergency room physician as a diagnostic tool on patients admitted with acute chest pain to guide the referral of patients either to the Coronary Care Unit or general ward | No control group |
| Ajay 2016[67] | Canada | Pre-post study | 20 | 5 government Community Health Centers in state of Himachal Pradesh, India | 5 clinics, 15-20 providers, 6797 patients | CVD management | Multicomponent intervention consisting of nurse care coordinator, structured training of the Medical Officers, mobile based CDS to generate individualized prescriptions for hypertension and diabetes, counseling services, follow-up care plan | No control group |

| Study ID | Country in which the study conducted | Study design | Duration of study (months) | Study setting | Sample size | Clinical domain of intervention | Intervention group | Control group |
|---|---|---|---|---|---|---|---|---|
| Mills 2018[68] | United States | Pre-post study | 50 | 3 urban EDs affiliated with one health system | 3 clinics, 558795 patients | CVD diagnostics | CDS with alerts based on Wells criteria for ordering CTPE | No control group |
| Shelley 2011[69] | United States | Pre-post study | 36 | A 4-site federally qualified CHC, Open Door Family Medical Centers in New York, USA | 5607 patients | Screening of CVD risk factors | CDS with guideline-based alerts and clinical reminders | No control group |
| Orchard 2019[70] | Australia | Prospective cohort | 20 | General practices across Sydney, Australia | 1805 patients | CVD management | CDS with guideline-based reminders for AF management | No control group |
| Lin 2013[71] | United States | Prospective cohort | 7 | Multicenter study | 100 providers, 472 patients | CVD diagnostics | CDS with guideline-based reminders to guide utilization of noninvasive imaging for individuals with suspected coronary artery disease | No control group |
| Pandya 2018[72] | Australia | Prospective cohort | 24 | 1 large metropolitan hospital and 1regional hospital in New South Wales, Australia | 205 patients | CVD management | CDS with guideline-based reminders for AF management | No control group |
| Fonarow 2010[73] | United States | Prospective cohort | 24 | 167 US outpatient cardiology practices. Community and academic cardiology or multispecialty outpatient practices | 167 cardiology practices, 34810 patients | CVD management | CDS with guideline-based management reminders for HF. Interventions included clinical decision support tools, structured improvement strategies, and chart audits with feedback. | No control group |
| Patel 2017[74] | Australia | QI intervention after completion of trial | 24 | 41 primary care clinics in Australia | 41 clinics, 22809 patients | CVD management | Multicomponent Health Tracker intervention consisting of CDS for risk assessment and management guidance, clinical audit tool, and performance report | No control group |
| Kim 2016[75] | United States | Retrospective study | 5 | A tertiary care center | 104 patients | Preventive care of CVD | CDS used by admitting service with guideline-based recommendations for risk assessment of VTE and initiation of prophylaxis | Assessment by consult service |
| Vecchio 2018[76] | Argentina | Retrospective study | 26 | A single center study in Argentina | 157 patients | Preventive care of CVD | CDS with guideline-based suggestions for treatment and diagnosis | No control group |
| Oppenkowski 2003[77] | United Kingdom | Retrospective study | 12 | 10 primary care centers in UK | 10 primary care centers, 367 patients | CVD management | CDS with guideline-based reminders for Warfarin initiation | No control group |

Abbreviation: RCT: randomized controlled trial; CVD: cardiovascular disease; CDS: clinical decision support; EHR: electronic health records; BP: blood pressure; AF: atrial fibrillation; HF: heart failure; PE: pulmonary embolism; ED: emergency department.

### eTable 2. Effectiveness of CDS, barriers to implementation, and strategies to improve implementation in 58 randomized trials

| Study ID | Outcomes measured | Effect on process outcome | Effect on clinical outcome | Economic outcome measured | Health disparity outcome measured | Unintended consequence measured or discussed | User experience (reported, Good/Neutral/Poor) | Directly test features associated of CDS effectiveness | Barriers of CDS implementation discussed | Strategies used to improve CDS implementation |
|---|---|---|---|---|---|---|---|---|---|---|
| Dregan 2014[1] | Last measure of blood pressure, cholesterol, prescribing for secondary prevention, cost of intervention | No effect | No effect | Yes | | | | | Lack of provider awareness/training | |
| Mazzaglia 2016[2] | Proportion of patients prescribed with cardiovascular drugs and days of drug–drug interaction exposure | Positive effect | | | | Alert fatigue | | | Alert fatigue | |
| Lester 2006[3] | Changes in hyperlipidemia prescriptions, time to prescription change, and changes in LDL levels | Positive effect | No effect | | | Alert fatigue | | | Alert fatigue | Repeated email communication with patients |
| Montgomery 2000[4] | Percentage of patients in each group with a five-year cardiovascular risk >10%, systolic blood pressure, diastolic blood pressure, prescribing of cardiovascular drugs. | No effect | No effect | | | Distraction | | | Alert fatigue, distraction | |
| Panjasawatwong 2015[5] | Time to return to SBP ≥80 mmHg, time until SBP remained ≥80 mmHg for at least 10 minutes, duration of hospitalization | | No effect | | | Alert fatigue | | | Alert fatigue, lack of useful information provided by the alerts | |
| Karlsson 2018[6] | Proportion of patients eligible for stroke prophylaxis who were prescribed anticoagulant therapy at 12 months, incidence of stroke, transient ischemic attack, or systemic thromboembolism, incidence of adverse event (bleeding) | Positive effect | No effect | | | Alert fatigue | Yes, will publish in a separate paper | | Alert fatigue, provider fear of stigma or doing harm | Integrate CDS with clinical workflow, deliver decision support at the time and location of decision making, provide actionable recommendations |
| Eckman 2016[7] | Proportion of patients with antithrombotic therapy that was discordant from CDS recommendation | No effect | | | | Alert fatigue, distraction | | | Complexity of real-world clinical situations and management of CVD, Alert fatigue | Integrate CDS with clinical workflow, embed CDS into EHR |
| Bucher 2010[8] | Total cholesterol, SBP, DBP and Framingham risk score | | No effect | | | | | Yes | Complexity of real-world clinical situations and management of CVD, data collection and follow-up of patients was not strictly endorsed | Integrate CDS with clinical workflow |
| Weir 2003[9] | Risk reduction in ischemic and hemorrhagic vascular events achieved by long-term anti-thrombotic therapy, and the proportions of subjects prescribed the optimal therapy identified by the CDS | No effect | No effect | | | | | | Discordance between local guidelines and CDS recommendations | |
| Keeffe 2005[10] | Providers' responses to computer-generated reminders for treatment of chronic heart failure | No effect | | | | | | Yes | Discrepancy between the intentions in the clinical guidelines and clinical practice | |

| Study ID | Outcomes measured | Effect on process outcome | Effect on clinical outcome | Economic outcome measured | Health disparity outcome measured | Unintended consequence measured or discussed | User experience (reported, Good/Neutral/Poor) | Directly test features associated of CDS effectiveness | Barriers of CDS implementation discussed | Strategies used to improve CDS implementation |
|---|---|---|---|---|---|---|---|---|---|---|
| Gill 2009[11] | Proportion of participants who were tested for hyperlipidemia, at lipid goal, and on lipid-lowering medications if not at goal | Positive effect | No effect | | | | | | High baseline rate of outcomes, lack of financial incentive, CDS not included patients with gaps in care | |
| Beeler 2014[12] | Prophylaxis orders placed 6–24 h after admission and transfer | Positive effect | | | | Reduction in alert fatigue, no increase in bleeding | | | Identification of individual risk factors by computers unreliable | Postponing prophylaxis checks to reduce number of reminders by 62% and minimize the risk of alert fatigue. |
| Luitjes 2018[13] | A combined rate of major maternal complications, guideline adherence, and patient-related outcomes | No effect | No effect | | | | Yes, good | | Lack of CDS integration into the workflow | Audit and provide feedback |
| Frances 2001[14] | Proportion of patients who had an active prescription for aspirin; proportion of patients with myocardial infarction who had an active -blocker prescription; proportion of patients receiving a cholesterol-lowering agent; and proportion of patients with a level of LDL cholesterol in the desired range (< 100 mg/dL). Secondary outcome variables included the proportion of patients who were hospitalized for myocardial infarction and patient mortality | No effect | No effect | | | | | | Lack of provider buy-in, static system without updates | |
| Hetlevik 1999[15] | Group differences in level of systolic and diastolic blood pressure, serum cholesterol, body mass index, and risk score for myocardial hypertension as well as group differences in fractions of smokers | | Mixed effect | | | | | | NA | |
| Kharbanda 2018[16] | Recognition of hypertension, guideline-adherent management, provider satisfaction | Mixed effect | | | | | Yes, good | | NA | Integrate CDS with clinical workflow, using precise triggers to reduce alert fatigue, audit and provide feedback, conduct ongoing training |
| O'Connor 2014[17] | BP and lipid control | | No effect | | | | | | NA | |
| Ranta 2015[18] | Guideline adherence and 90-day stroke risk, cerebrovascular/vascular/death/adverse events, cost, and user feedback | Positive effect | Mixed effect | Yes | | high number of unintended registrations | Yes, good | | Provider fear of stigma or doing harm | |
| Nendaz 2010[19] | Difference in the adequacy of thromboprophylaxis prescription between the baseline survey and the post-CDS implementation assessment | No effect | | | | Improvement in the rate of over prescription with alerts. | | | Providers distrust of CDS recommendations, lack of provider training, discordance between local guidelines/policies and CDS recommendations | Obtain support from the local leader followed by reminders during the intervention period |

| Study ID | Outcomes measured | Effect on process outcome | Effect on clinical outcome | Economic outcome measured | Health disparity outcome measured | Unintended consequence measured or discussed | User experience (reported, Good/Neutral/Poor) | Directly test features associated of CDS effectiveness | Barriers of CDS implementation discussed | Strategies used to improve CDS implementation |
|---|---|---|---|---|---|---|---|---|---|---|
| Sperl-Hillen 2018[20] | Predicted 1-year CVD risk | Positive effect | | | | | Yes, good | | Technical problems with CDS or EHR infrastructure | Integrate CDS with clinical workflow, limit CDS display and print burden, conduct ongoing training, audit and provide feedback, trigger CDS by rooming staff rather than PCPs, provide financial incentives |
| Chaturvedi 2019[21] | OAC utilization | No effect | | | | | | | Technical problems with CDS or EHR infrastructure | |
| Roy 2016[22] | A composite endpoint comprising of symptomatic VTE events and major bleedings during the three months following hospital admission, VTE events, major bleedings, and all-cause mortality during the 3 months | No effect | No effect | | | | | | Technical problems with CDS or EHR infrastructure, complexity of real-world clinical situations and management of CVD | |
| Fitzmaurice 2000[23] | Therapeutic control of the international normalized ratio, intervention cost | No effect | | Yes | | | Yes, good | | Technical problems with CDS or EHR infrastructure, time and resource constraints | |
| Holbrook 2011[24] | The primary outcome was a composite score of 8 recommended process outcomes at 1 year. The main secondary outcome was a clinical composite score based on the same 8 risk factors | Positive effect | No effect | | | | | | Technical problems with CDS or EHR system | Improve technical integration with EHR |
| Gilutz 2009[25] | Compliance to treatment, LDL levels, Rate of cardiac hospitalizations | Positive effect | Positive effect | | | | | | Time and resource constraints | Use precise triggers to reduce alert fatigue |
| Arts 2017[26] | Acceptance of CDS alert, clinical improvement for stroke prevention in AF patients | No effect | No effect | | | Alert fatigue | Yes, neutral | Yes | Time and resource constraints, alert fatigue, technical problem with CDS, Alert fatigue | Alert prioritization, user customization, tight EHR integration, strict selection of alerts |
| vanEngen-Verheul 2015[27] | Guideline concordance to national CR guidelines | No effect | | | | | | | Time and resource constraints, discordance between local guidelines and CDS recommendations | Audit and provide feedback, use iterative cycles based on quarterly feedback reports to improve implementation |
| Hetlevik 2000[28] | Group differences in fractions of patients without registrations of CVD risk factors (process evaluation) and mean group differences for the same variables (patient outcome evaluation). | No effect | No effect | | | | Yes, poor | | Time and resource constraints, discrepancy between the intentions in the clinical guidelines and clinical practice | |
| Wells 2017[29] | Proportion of patients with a completed CVD risk assessment | No effect | | | | | Yes, poor | | Time and resource constraints, lack of CDS integration into the workflow | |

| Study ID | Outcomes measured | Effect on process outcome | Effect on clinical outcome | Economic outcome measured | Health disparity outcome measured | Unintended consequence measured or discussed | User experience (reported, Good/Neutral/Poor) | Directly test features associated of CDS effectiveness | Barriers of CDS implementation discussed | Strategies used to improve CDS implementation |
|---|---|---|---|---|---|---|---|---|---|---|
| Eccles 2002[30] | Adherence to the guidelines, based on review of case notes and patient reported generic and condition specific outcome measures | No effect | No effect | | | | | | Time and resource constraints, lack of CDS integration into the workflow, complexity of real-world clinical management of CVD. | |
| Cheng 2018[31] | Change in SBP, LDL reduction, antithrombotic medication use, smoking cessation, and physical activity | | Mixed effect | | | | | | Time and resource constraints, lack of provider buy-in | Establish EHR systems to standardize collection of data and monitor adherence to intervention protocols and integrate CDS with EHR |
| vanDoorn 2018[32] | A composite of stroke, TIA and/or thromboembolism, bleeding and the proportion of patients on guideline recommended anticoagulant treatment | No effect | No effect | | | Alert fatigue | | | Time and resource constraints, perceived lack of added value to the workflow, Alert fatigue | |
| Subramanian 2004[33] | Physician adherence to heart failure guidelines, as well as patients' NYHA class, quality of life, satisfaction with care | Mixed effect | No effect | | | | | Yes | Time and resource constraints, providers distrust CDS data | |
| Anchala 2015[34] | Change in blood pressure at 12 months, cost-effectiveness ratio, physician performance and in quality of care given for patients | Positive effect | Positive effect | Yes | | | | | | Integrate CDS with clinical workflow, conduct ongoing training, engage stakeholder in CDS development process, multicomponent intervention with concurrent advice to physicians and patients in institutional settings |
| Roy 2009[35] | Appropriateness of diagnostic work-up, adherence to guideline recommendations (secondary outcome); number of tests per patient (secondary outcome). | Positive effect | | | | | | | | |
| Tian 2015[36] | Proportion of patient-reported antihypertensive medication use from baseline to 1-year follow-up. Secondary outcomes: (1) proportion of high-risk individuals taking aspirin, (2) mean systolic blood pressures of high-risk individuals, (3) proportion of current smokers, (4) proportion of high-risk individuals aware of the harms of a high-salt diet, (5) the proportion of high-risk individuals receiving monthly follow-ups from the community health workers, and (6) proportion of high-risk individuals hospitalized | Positive effect | No effect | | | | | | | Cultural tailoring of lifestyle interventions |

| Study ID | Outcomes measured | Effect on process outcome | Effect on clinical outcome | Economic outcome measured | Health disparity outcome measured | Unintended consequence measured or discussed | User experience (reported, Good/Neutral/Poor) | Directly test features associated of CDS effectiveness | Barriers of CDS implementation discussed | Strategies used to improve CDS implementation |
|---|---|---|---|---|---|---|---|---|---|---|
| Guo 2017[37] | Patients' knowledge, quality of life, drug adherence, and anticoagulation satisfaction, Usability, Feasibility, Acceptability of CDS | Positive effect | | | | | Yes, good | | | |
| Goud 2009[38] | Concordance with guideline recommendations | Positive effect | | | | | | | | Conduct local consensus discussions |
| B. Sussman 2018[39] | Prescription of a medium- or high-strength statin, retention of the intervention's effects after it was completed | Positive effect | | | | | | | | Integrate CDS with clinical workflow and EHR |
| Cleveringa 2008[40] | 1-year difference in A1C, 1-year difference in the 10-year UKPDS CHD risk estimate and the percentage of patients that reached A1C >=7%, systolic blood pressure >=140 mmHg, total cholesterol>=4.5 mmol/l, and LDL cholesterol>=2.5 mmol/l, cost-effectiveness ratios | | Mixed effect | Yes | | | | | | |
| Peiris 2015[41] | (1) guideline-indicated risk factor measurements and (2) guideline-indicated medications for those at high cardiovascular disease risk. | Mixed effect | | | Yes | | | | | Integrate CDS with clinical workflow, alignment with usual decision-making processes in the patient consultation, provision of treatment recommendations rather than just assessments, audit and feedback |
| Peiris 2019[42] | Proportion meeting systolic blood pressure (SBP) targets (<140mmHg) | | No effect | | | High discordance rate between the people identified at high CVD risk in the evaluation dataset and those identified by community workers, a higher-than-expected improvement in BP treatment rates in the control period | Yes, good | | | Integrate CDS with clinical workflow |
| Ali 2016[43] | Proportion of patients achieving HbA1c<7.0% and BP<130/80mmHg or/and LDL cholesterol <100mg/dl (primary outcome); mean risk factor reductions, health-related quality of life, and treatment satisfaction (secondary outcomes) | Positive effect | Positive effect | | | | Yes, good | | | Obtain local 'buy-in' at pre-adoption stage, show evidential benefit of CDS |
| Johnston 2019[44] | Proportion of patients with a favorable outcome based on the 90-day modified Rankin Scale score | | No effect | | | | | | | |
| Zhu 2018[45] | Blood pressure, self-care behaviors, self-efficacy, quality of life and satisfaction | Positive effect | Positive effect | | | | | | | |

| Study ID | Outcomes measured | Effect on process outcome | Effect on clinical outcome | Economic outcome measured | Health disparity outcome measured | Unintended consequence measured or discussed | User experience (reported, Good/Neutral/Poor) | Directly test features associated of CDS effectiveness | Barriers of CDS implementation discussed | Strategies used to improve CDS implementation |
|---|---|---|---|---|---|---|---|---|---|---|
| McKinstry 2013[46] | Mean daytime SBP, DBP | | Positive effect | | | | | | | |
| Hicks 2008[47] | BP Control and adherence to medication | Positive effect | No effect | | Yes | | | | | |
| Brocklehurst 2018[48] | Neonatal outcome, developmental assessment at age 2 years | No effect | No effect | | | | | | | |
| Piazza 2009[49] | VTE within 90 days | | No effect | | | | | | | Increasing resources for computer-based decision-support strategies and medical informatics |
| Kucher 2005[50] | Confirmed DVT or PE within 90 days | | Positive effect | | | | | | | Integrate CDS with clinical workflow and EHR |
| Bhavnani 2018[51] | Time to referral for therapy with percutaneous valvuloplasty or surgical valve replacement, probability of a cardiovascular hospitalization and/or death over 1 year | Positive effect | Positive effect | | | | | | | |
| Mahler 2015[52] | Objective cardiac testing (stress testing or angiography); Index length of stay, early discharge, and major adverse cardiac events | Positive effect | Positive effect | | | | | | | |
| Tutino 2017[53] | Proportion of patients achieving ≥ 2 treatment targets (HbA1c< 53 mmol/l (7%), blood pressure < 130/80 mmHg and LDL cholesterol < 2.6 mmol/l); Other outcomes included default rates, change in quality-of-life measures, frequency of hypoglycemia, adherence to lifestyle modification/self-care activities, and new onset of physician-documented diabetes-related endpoints | Positive effect | No effect | | | | | | | |
| Jackson 2012[54] | Changes in SBP, DBP | | Positive effect | | Yes | | | | | |
| Huebschmann 2012[55] | Clinical inertia and BP control | Positive effect | No effect | | | | | | | |
| Bosworth 2007[56] | BP Control | | Positive effect | | | | | | | |
| Smith 2008[57] | Diabetes care processes (diabetes test completion), outcomes (metabolic and cardiovascular risk factors, estimated coronary artery disease risk), and patient costs (payer perspective) | No effect | No effect | Yes | | | | | | |
| Kline 2014[58] | Reduction in the proportion of healthy patients exposed to >5 mSv chest radiation. | Positive effect | | | | | | | | |
